# Comparative effectiveness of first line pembrolizumab vs. chemotherapy in aNSCLC: A Norwegian Population-Based Cohort Study

**DOI:** 10.1101/2024.10.07.24314963

**Authors:** Simon Boge Brant, Siri Børø, Christian Jonasson, Åslaug Helland, Steinar Østerbø Thoresen

## Abstract

The KEYNOTE-042(1,2) trial showed a benefit of treating patients with non-oncogene addicted advanced NSCLC with PD-L1 tumour proportion score over 50% with Pembrolizumab as monotherapy over platinum doublet chemotherapy. To contextualize these results, we undertake a detailed emulation of the inclusion criteria in Keynote–042 using Norwegian health registry data, and discuss both the clinical context, as well as the general utility of such registry data for pharmacoepidemiologic research in oncology. Within the population of patients with PD-L1 tumour proportion score over 50%, an observational analogue of an intention to treat analysis showed similar results to those of the Keynote-042 study.

## 1 Introduction

The KEYNOTE-042 (KN-042) trial was published in 2019 and is a phase 3 RCT of Pembrolizumab monotherapy versus chemotherapy as first line treatment in patients with locally advanced or metastatic non-small-cell lung cancer with a PD-L1 Tumour Proportion Score (TPS) of 1% or greater (1). For KN-042 (1,2), there are several relevant observational studies who have investigated the RW-effectiveness of Pembrolizumab but to our knowledge there are no studies that focus on overall survival in a nationwide registry setting, within a European context. The purpose of this study was to study the effectiveness of Pembrolizumab compared to platinum-based chemotherapy in a real-world (RW), Norwegian, population-based health data-cohort, by using a target trial emulation approach (3,4) based on the eligibility criteria of the KN-042 trial. In addition to their clinical relevance, our results highlight the potential usefulness of observational data from Nordic health registries in drug development, such as conducting externally controlled studies or registry-embedded clinical trials (5).

## 2 Material and Methods

### Data sources

We used nationwide data from three population-based registries in Norway: The Cancer Registry (CRN), the National Patient Registry (NPR) and the Norwegian prescribed drug registry (NorPD). The data sources contains comprehensive overview of cancer specific diagnostics, treatment modalities (surgery, radiation, and medical treatment) and follow-up, information on all patients who have been referred to or have received specialized healthcare at any specialist health-care service in Norway with ICD-10 codes for diagnostic data, administrative -, demographic-, and reimbursement information, as well as information on dispensed drugs (by prescription) from pharmacies. These registries have mandatory reporting without consent. The unique, 11-digit personal identification number (PIN) each citizen living in Norway has, allows for linkage between them. For a more detailed description, see (6–9).

Patient-level data were de-identified to guarantee the protection of individual patient integrity, and ethical approval was obtained from the Regional Committee for Medical and Health Research Ethics, South-East D (ID number 108024), which waived the requirement of informed consent.

### Study design

Patients were selected from a cohort of all patients diagnosed with lung cancer in Norway in the period 1st of January 2017 – 30th of September 2021, for whom follow up information was available up to the 31st of December 2021, and whose treatment in the first line was either A: Pembrolizumab alone, or B: chemotherapy. Chemotherapy was defined as at least one platinum – based drug, plus at least one other second or third generation chemotherapy agent such as Paclitaxel, Vinorelbine, or Etoposide. First line treatment was defined as all the treatment (drugs, resection, and radiotherapy) the patient received starting from the first treatment after a maximum of 30 days before the lung cancer diagnosis, until 30 days after the first treatment the patient received. Patients who received radiotherapy or surgical treatment in the first line were not considered.

Figure 1 describes the patient selection process. From the initial patient pool, patients were removed if they did not conform to a list of criteria, referred to as C1 to C12. These criteria are the same eligibility criteria defined in the KN-042 trial. To be included, the patients needed to have advanced or metastatic lung cancer with non-small cell histology, not be ALK or EGFR positive, have ECOG status 0 or 1, have adequate organ function, have had no other primary cancer the last 5 years, not have received paclitaxel + carboplatin in the adjuvant setting if they have squamous cell lung cancer, not have active CNS metastases, not have active hepatitis B or C, be HIV negative, not have any active autoimmune disease that has required treatment for two years, not have received prior systemic chemotherapy treatment for lung cancer, have had no prior biological therapy, not have had major surgery for three weeks, not have received radiation therapy (except with palliative intent) for 6 months, and not have any prior exposure to anti PD-L1 / PD-1 drugs.

**Figure 1:**
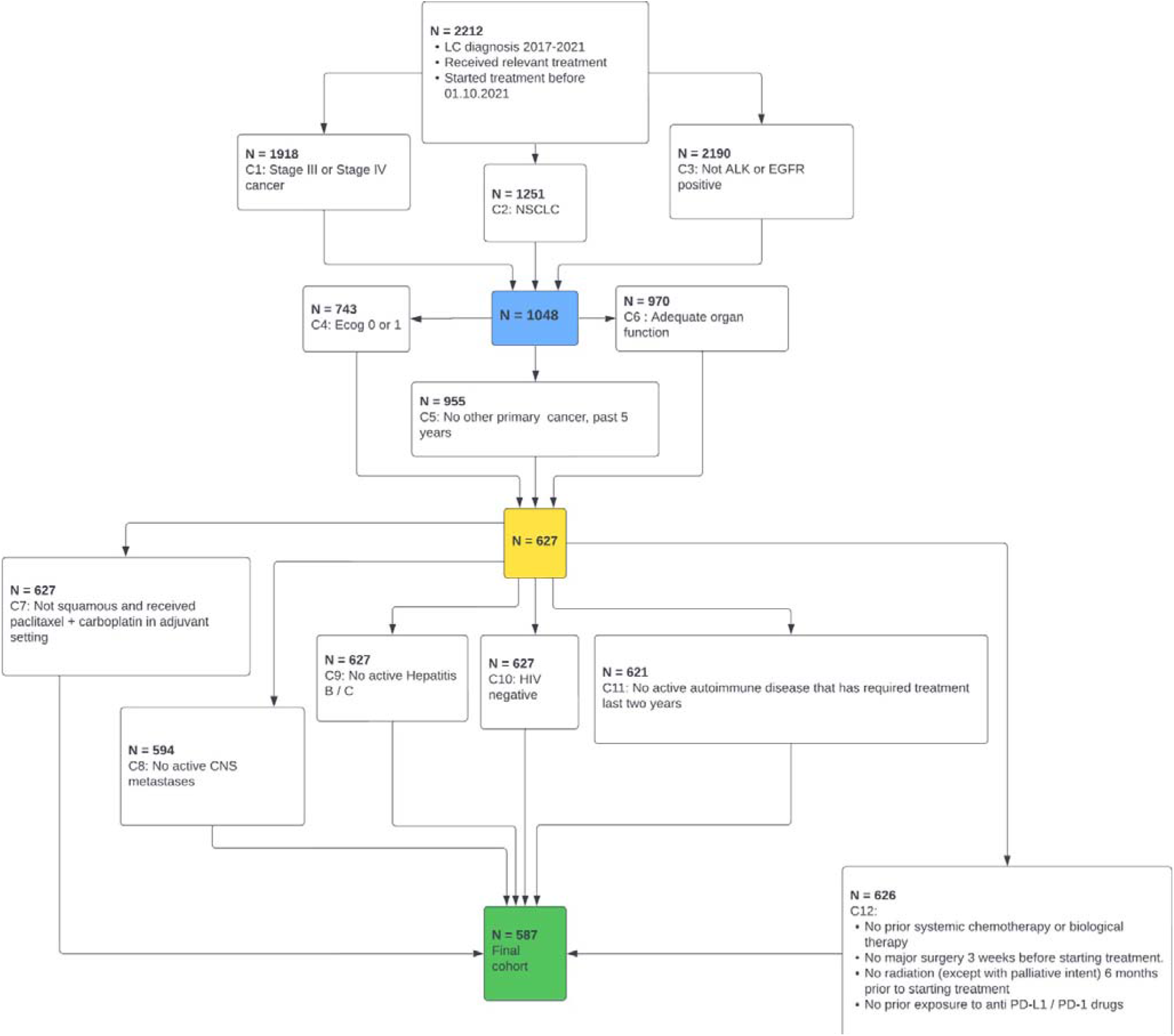
Patient selection, through each 11 eligibility criteria (C1-C12).

Adequate organ function was defined as that the patient had not been given an ICD-10 diagnosis in the past year that indicated either neutropenia, thrombocytopenia, acute myocardial infarction, heart failure, moderate to severe kidney disease, or severe liver disease. Patients were considered to have active CNS metastases if they had been given either of the ICD-10 diagnoses C793 or C794 in the last four weeks leading up to the initiation of the treatment. The final cohort of patients consisted of 587 patients, where 345 and 242 had been treated with chemotherapy and pembrolizumab in the first line, respectively.

### Outcome definitions

As outcomes, we consider overall survival from the initiation of the first line treatment, as well as an amended version where survival times were censored at the time the patient switched to a second-line treatment if this happened before the patients completed 4 courses of a platinum drug, or 35 courses of Pembrolizumab, depending on which treatment they initiated. The analysis of the outcome where artificial censoring is introduced when patients switched treatment in these cases is referred to as *per protocol*, or *PP*, whereas the analysis of the outcome without this censoring is referred to as *intention to treat*, or *ITT*.

### Statistical analysis

Propensity score weighting (inverse probability of treatment weighting) was used to attempt to minimize confounding bias, using the patients ECOG status, stage and histology at diagnosis, age, gender, and Charlson comorbidity score. Differences in outcomes among the two treatment arms in both the ITT-, and PP-analysis were assessed by examining Kaplan-Meier curves and hazard ratios, with and without weighting. In the weighted analyses, confidence intervals are computed by bootstrapping.

## 3 Results

The patient characteristics are summarized in Table 1. The patient groups having received chemotherapy or Pembrolizumab as their 1L treatment were balanced for age, ECOG -status, metastatic stage, and squamous histology stage. The proportion of males were larger in the chemotherapy group compared to the Pembrolizumab group, respectively 57.7 % vs 45.9 %. We report the PD-L1 status but note that we did not take the patients PD-L1 status into account when defining the patient population, or in the weighted analysis. This should be kept in mind when interpreting these results and it is elaborated under the discussion section.

**Table 1:**
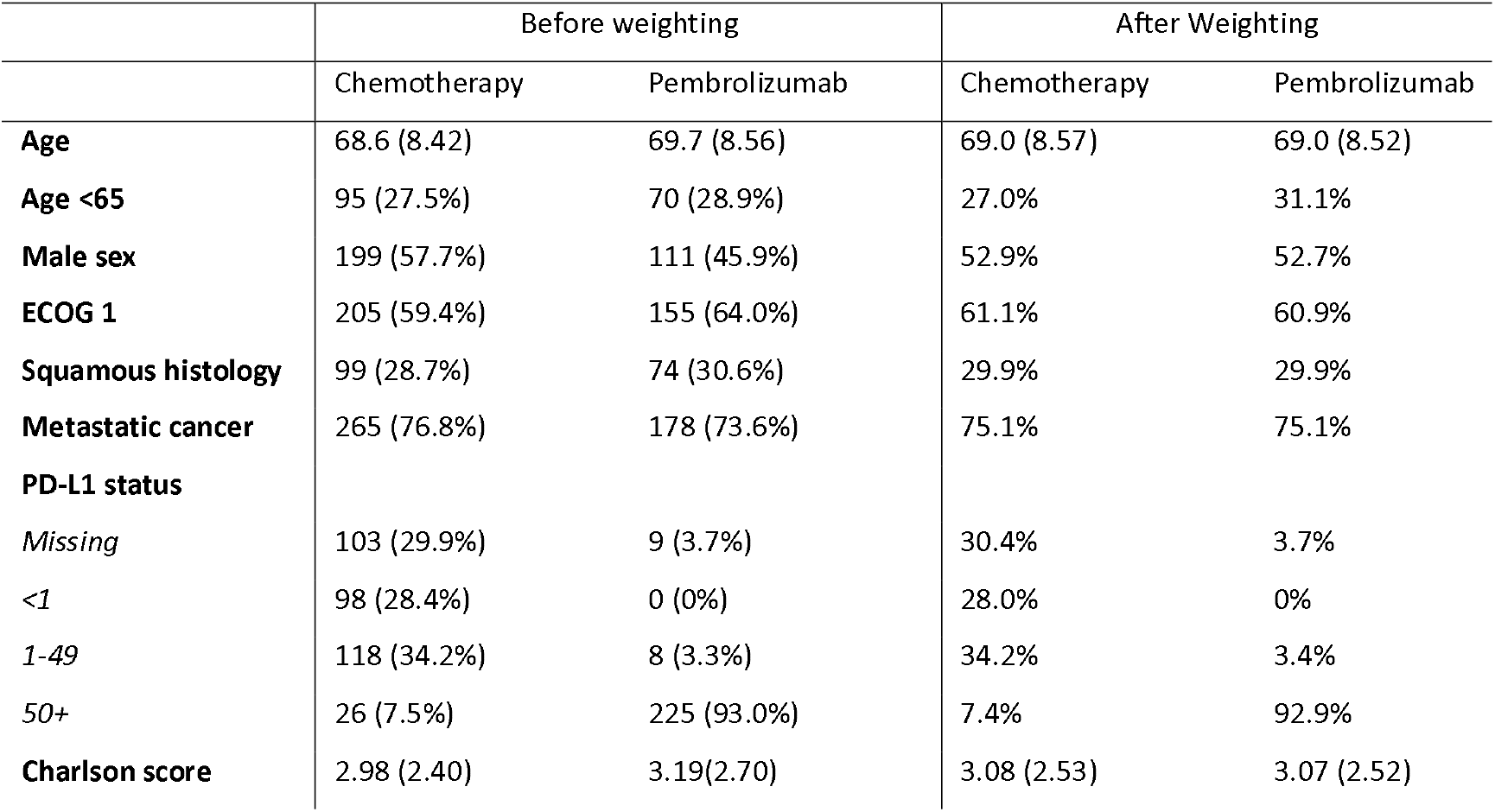
Baseline characteristics, with and without propensity score weighting (weighting variables: ECOG status, stage and histology at diagnosis, age, gender, and Charlson comorbidity score)

Overall, compared with chemotherapy as first line treatment, real-world patients treated with Pembrolizumab in the first line had a longer survival in both the ITT-, and PP-analysis. Kaplan Meier curves and hazard ratios (HR) are shown in Figure 2 for both outcomes, with and without weighting. For the ITT outcome, the median survival was 21 months [95%CI 17-27] vs 10 months [95%CI 9-12], for respectively Pembrolizumab vs chemotherapy, with a HR of 0.56 (0.45, 0.68). The median follow-up time was 11 months (10 months for the chemotherapy group, and 15 months for the pembrolizumab group). For the PP outcome, the median survival was 34 months [95%CI 25-NE] vs 11 months [95%CI 9-13] for Pembrolizumab vs chemotherapy with a HR of 0.44 (0.34, 0.57). The median follow-up time was 8 months (8 months for the chemotherapy group, and 10.5 months for the pembrolizumab group). Propensity score weighting the study arms in the two groups did not change the superiority of Pembrolizumab vs chemotherapy in the two groups, respectively 21 months [95%CI 18-27] vs 10 months [95%CI 9-12] with a HR of 0.44 (0.34, 0.57) for the ITT-analysis, while the weighted HR was 0.42 (0.32, 0.54) for the PP-analysis with a median survival of 34 months [95%CI 25-NE] vs 11 months [95%CI 9-12] for respectively Pembrolizumab vs chemotherapy.

**Figure 2:**
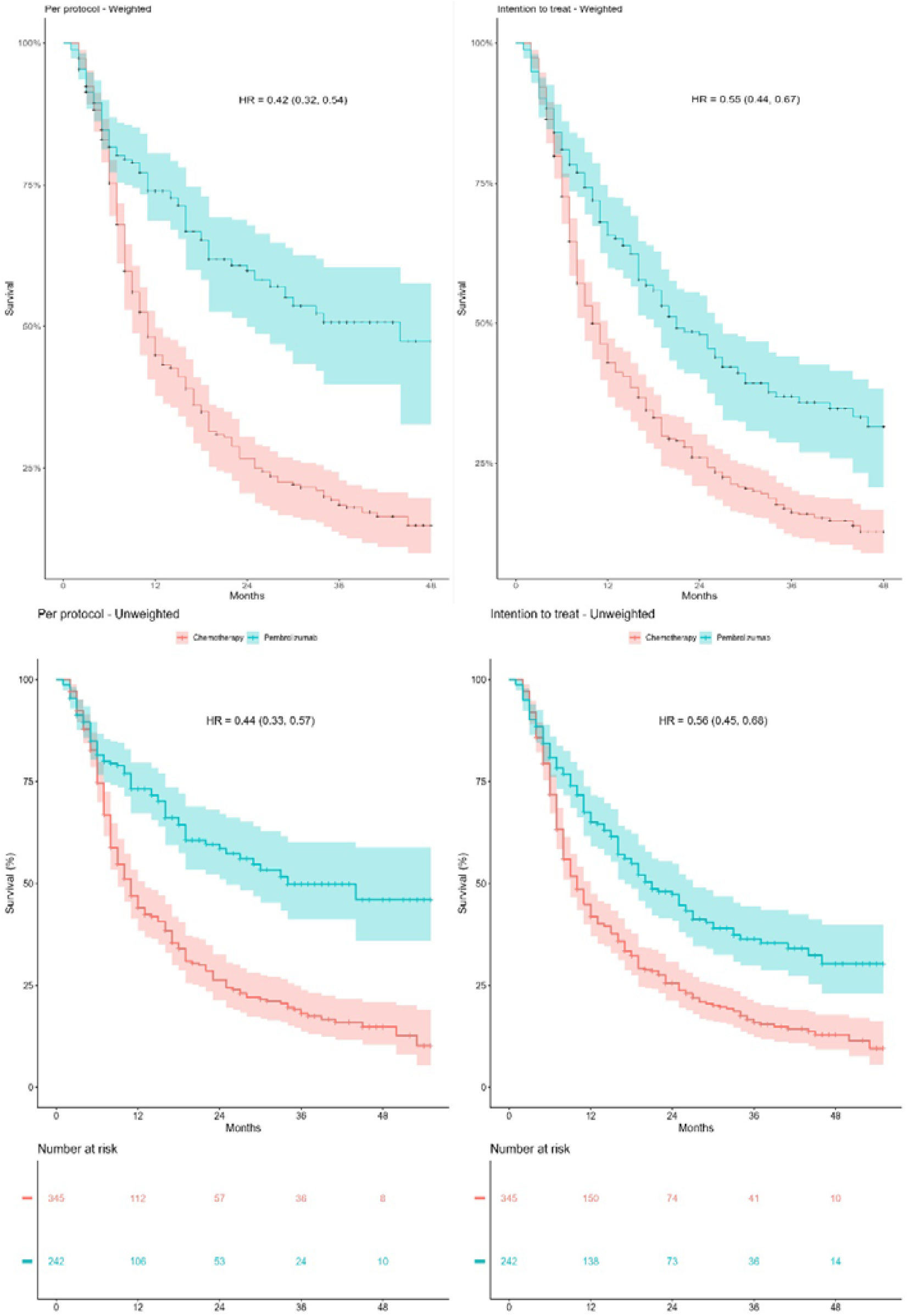
KM-curves for overall survival in the ITT-, and PP-group, with and without propensity score weighting (weighting variables: ECOG status, stage and histology at diagnosis, age, gender, and Charlson comorbidity score).

**Figure 3:**
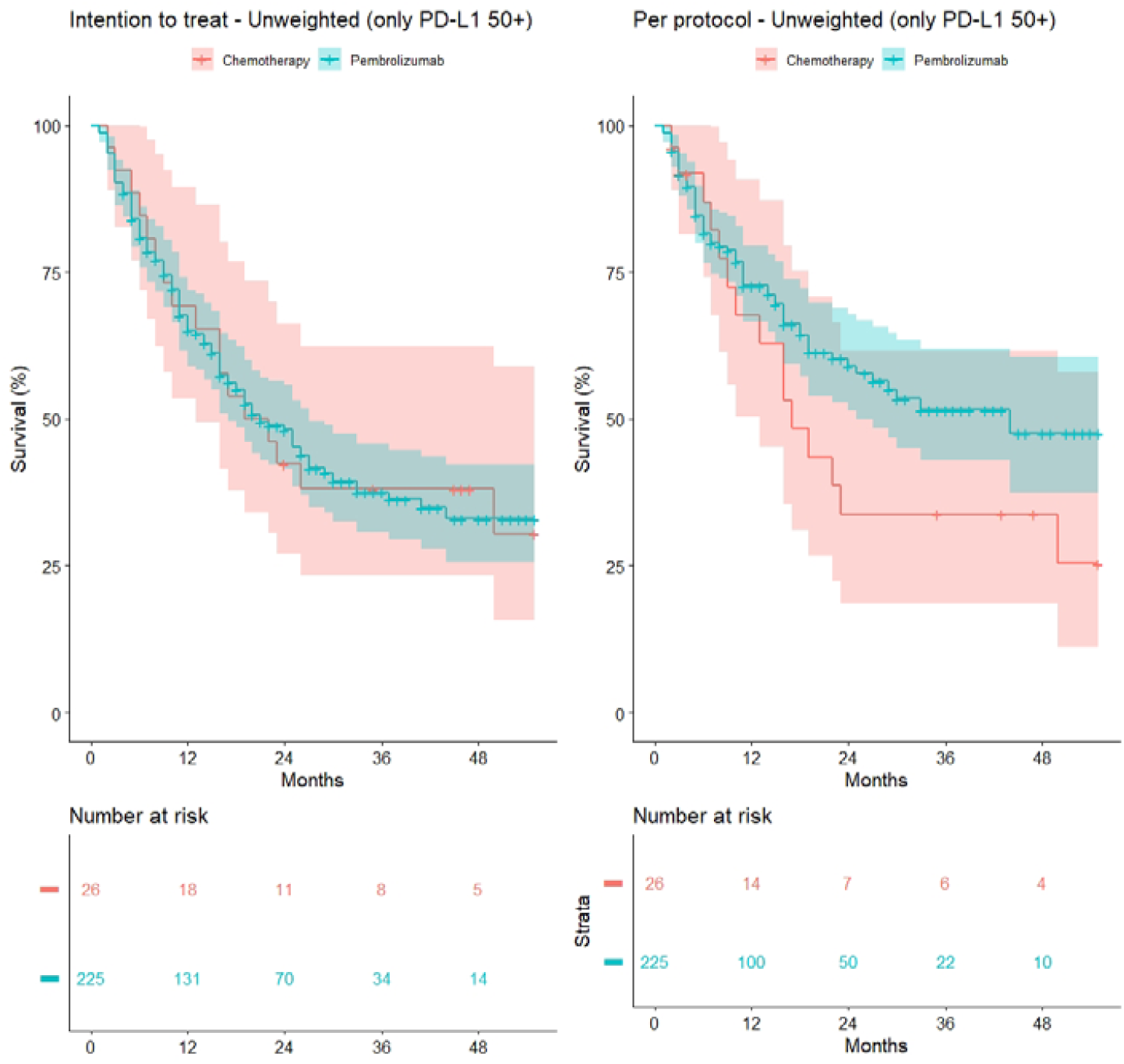
Unadjusted KM curves for patients who have a recorded PD-L1 expression of over 50%.

There was a maximum 4-year follow-up period in the data. The differences in the 1–4-year survival between the weighted and the unweighted groups, were small for both the survival outcomes (see Table 2). With the exception of the 1-year survival of the chemotherapy patients, the 95% confidence interval for the 1–4-year survival all contained the corresponding x-year survival reported in KN-042 for the patients with TPS over 50%. The RCT OS-data are consistently higher than the RW OS-data for the chemotherapy arm while it is opposite for the Pembrolizumab arm, where the RW OS-data are consistently higher than the RCT OS-data, exemplified by the 4-year survival data of the patients in KN-042 which were 13.5 % and 25.3% in respectively the chemotherapy arm vs Pembrolizumab arm and the RW cohort had a 4-year survival of respectively 12.8% and 29.6%.

**Table 2:** Overview of median survival, as well as 1-, 2-, 3-, and 4-year survival data in both the ITT-, and PP-group. Results for the propensity score weighted analysis are also included. Corresponding survival data from KN-042 included for reference.

| Intention to treat |  |  |  |  |  |  |
| --- | --- | --- | --- | --- | --- | --- |
| Parameter | Chemotherapy RWE | Chemotherapy RWE (weighted) | Pembrolizumab RWE | Pembrolizumab RWE (weighted) | Chemotherapy RCT | Pembrolizumab RCT (4, 11) |
| Median (months) | 10 (9, 12) | 10 (9, 12) | 21 (17, 27) | 21 (18, 27) | 12.2 (10.4, 14.2) | 20 (15.4, 24.9) |
| 1-year survival | 41.9% (37.0, 47.5) | 42.2% (36.8, 47.3) | 65.0% (59.1, 71.6) | 66.4% (59.9, 72.5) | 50.7% | 63.5% |
| 2-year survival | 25.5% (21.1, 30.7) | 25.4% (21.0, 30.2) | 47.3% (40.8, 54.8) | 47.7% (40.0, 54.9) | 29.9% | 44.5% |
| 3-year survival | 15.9% (12.2, 20.6) | 15.7% (11.9, 20.0) | 36.4% (29.8, 44.5) | 36.2% (28.7, 44.1) | 18.4% | 31.3% |
| 4-year survival | 12.9% (9.3, 17.8) | 12.8% (8.9, 16.8) | 30.3% (23.0, 39.8) | 29.6% (20.7, 38.3) | 13.5% | 25.3% |
| Per protocol |  |  |  |  |  |  |
| Median (months) | 11 (9, 13) | 11 (9, 12) | 34 (25, -) | 34 (25, -) | - | - |
| 1-year survival | 44.0% (38.4, 50.5) | 43.8% (37.8, 49.7) | 71.1% (67.2, 79.6) | 74.9% (68.6, 80.5) | - | - |
| 2-year survival | 26.3% (21.3, 32.5) | 25.8% (20.6, 30.6) | 58.5% (51.0, 67.1) | 59.1% (49.9, 67.9) | - | - |
| 3-year survival | 18.0% (13.6, 23.8) | 17.8% (13.1, 22.8) | 49.8% (41.2, 60.2) | 50.5% (39.9, 60.4) | - | - |
| 4-year survival | 14.8% (10.5, 20.9) | - | 46.0% (36.0, 58.8) | - | - | - |

## Discussion

In this paper we have assessed the comparative effectiveness of Pembrolizumab and platinum-based chemotherapy using a target trial emulation approach based on the eligibility criteria of the KN-042 trial, using observational data from Norwegian health registries. We considered overall survival for an observational analogue of intention to treat (ITT), by identifying patients who initiated a treatment consistent with one of the two treatments in the KN-042 trial. In an RCT, the ITT effect is the effect of the being allocated to a treatment (10), whereas for the observational analogue, it is the effect of initiating a treatment. In addition, we also considered an observational analogue of a per-protocol (PP) analysis, where patients who initiated one of the treatments were censored if they switched treatment before completing the treatment plan as described in the KN-042 study.Introducing such a censoring mechanism will in most cases introduce dependent censoring. Therefore, we also explored using censoring weights based on the same variables as the treatment weights using the approach described by (11). However, this did not change the estimates noticeably, so in the analyses presented here estimates are without censoring weights.

The treatment effect of pembrolizumab compared with chemotherapy is larger in our dataset than in KEYNOTE-042. The per–year survival in our observational cohort is lower than the corresponding per-year survival from KEYNOTE-042 for the chemotherapy group, and higher than the corresponding per-year survival for the pembrolizumab group. Historically, an important discussion point of the KN-042 trial was that the design did not allow for crossover from the chemotherapy to the pembrolizumab arm (1), and it was questioned whether this may have caused the survival of the chemotherapy patients to be lower than what it would have been if they had received what was the standard of care during the study period. In our analysis, however, enforcing the protocol on the chemotherapy group by censoring patients that moved to a second line treatment before they completed 4 courses only increased the median survival by 1 month, and between 1-2 percentage points for each per-year survival. By comparison, the median survival in the chemotherapy arm of the CheckMate 026 study (12), where crossover to the nivolumab arm was allowed, was 13.2 months (10.7 – 17.1). Further, in earlier studies conducted before PD-L1 inhibitors were introduced for these patients, median overall survival for platinum-doublet chemotherapy treated patients ranged between 10 -13 months (13–15).

One should note that because the Keynote-042 study did not show significant benefit of using PD-L1 inhibitors over chemotherapy in patients with low PD-L1 tumour proportion score, monotherapy with pembrolizumab has not been used as first line treatment for these patients in Norway. Therefore, we can only attempt to replicate the results for the subgroup with a PD-L1 tumour proportion score of at least 50%. Dates of PD-L1 tests were not available, and there were some patients for whom a PD-L1 status was not recorded. Therefore, it was not possible for us to determine at which point in time patients who initiated chemotherapy and had a recorded PD-L1 status were tested. In addition, when comparing the reported 1–5-year survival from the Keynote 042 study (2) for the chemotherapy patients stratified by high/low PD-L1 status, survival after receiving chemotherapy treatment seems to be unaffected by PD-L1 status in the short term.

Therefore, it is natural to assume that any effect modification of effectiveness of the chemotherapy by PD-L1 status is mediated by the patients later treatment lines. Juxtaposed with the survival curves from our cohort (see Figure 4) these differences seem to be negligible. We therefore think that we can reasonably assume that for a patient who received pembrolizumab, their survival time would have followed the same distribution as a patient in who received chemotherapy who, apart from their PD-L1 status is otherwise identical, if they had been treated with chemotherapy. Based on this we chose not to include the patients PD-L1 score in the propensity score model, and interpret the effect estimate as being valid for a patient population that has the distribution of PD-L1 scores of the group of patients that received pembrolizumab. We have to the best of our ability tried to minimise confounding by controlling for known factors at time zero that are likely determinants of the patients’ outcome, such as their ECOG status, stage of disease, and comorbidities summarised by the Charlson score. However, we can only make adjustments based on available data, so there are some factors that we do not know the distribution of within the two groups that could be important, most notably smoking status. Despite these limitations, our analyses indicate a superiority of monotherapy with pembrolizumab over a platinum-doublet chemotherapy in this population, on a scale similar to those shown in Keynote-042, although the estimated effect is somewhat larger in the clinical setting. This could have several explanations, for instance that the estimand is subtly different (the effect of initiating a treatment versus the effect of being randomized to a treatment group), there could be differences in the distribution of patient characteristics that modify the effect of the treatments, and differences in second line treatments for those that progress. The level of detail of the data, and its apparent appropriateness for studying such treatment effects illustrates some of the utility that observational data from registries such as those from Norway can have, for instance for drug development via registry-embedded clinical trials (5).

**Figure 4:**
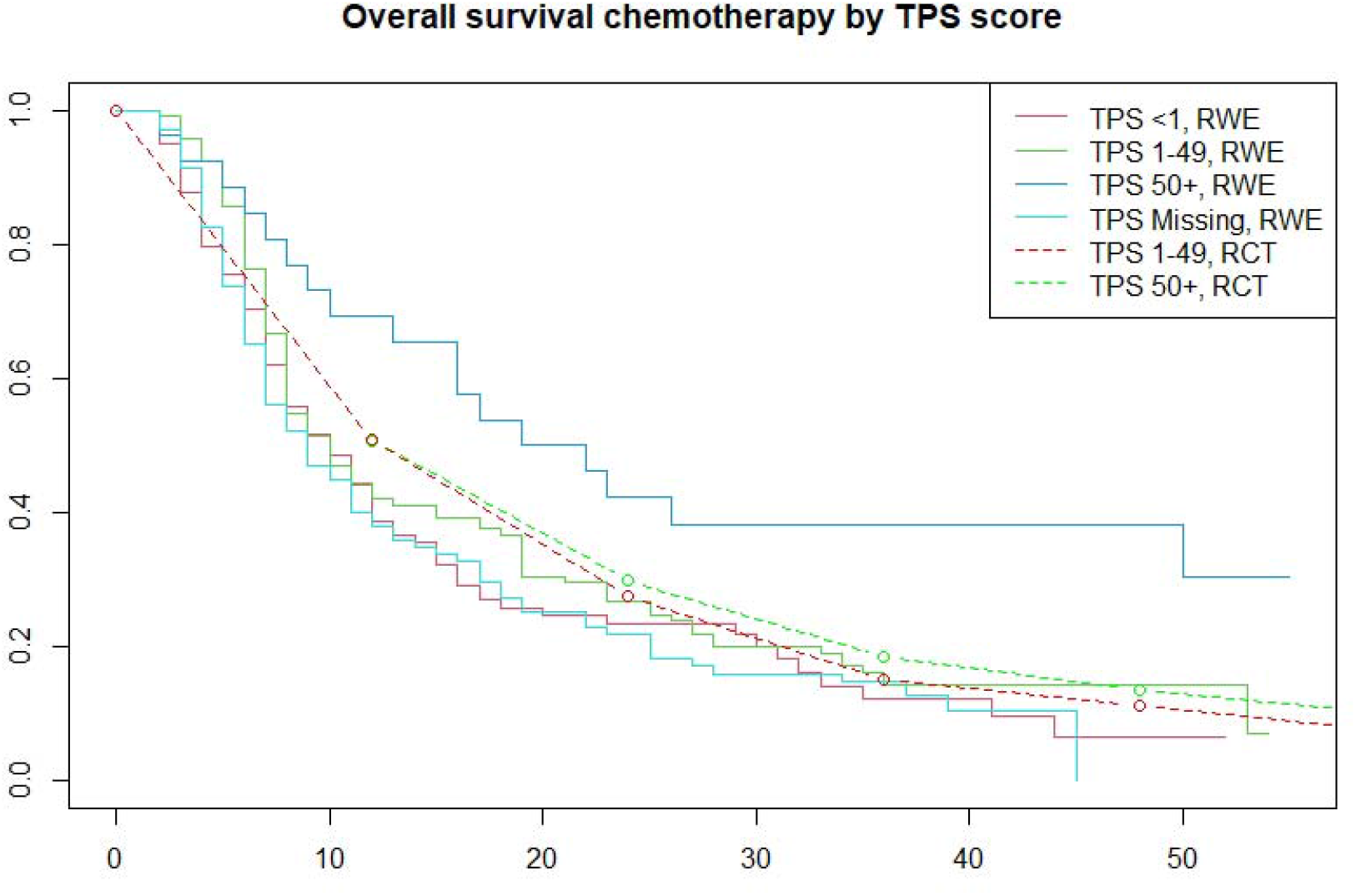
Comparison of OS within strata of PD-L1 score for 1L-chemotherapy.

## 5 Study Highlights

### What is the current knowledge on the topic?

There are some relevant observational studies who have investigated the RW-effectiveness of Pembrolizumab (16–18) but to our knowledge there are no studies that focus on overall survival in a nationwide registry setting, within a European context.

### What question did this study address?

Real-world comparative effectiveness between Pembrolizumab and platinum-based doublet chemotherapy, in a Norwegian population-based cohort, for a subgroup of NSCLC patients.

### What does this study add to our knowledge?

The real-world evidence literature that concerns extensive emulation of published randomized clinical trials is largely populated by studies based on US databases (19, 20). Utilizing Norwegian health data, thus represents a contribution to the diversification of real-world evidence sources, as one reduces the inherent issue of patient, -and treatment selection due to the mandatory reporting and population-based nature of the data sources. Further, it specifically illustrates the usefulness of Norwegian data for research purposes.

### How might this change clinical pharmacology or translational science?

Our study might change clinical pharmacology or translational science by highlighting and providing evidence of the usefulness of Nordic health registries, which could encourage more interest and investment in registry-based research.

## 6 Acknowledgements

## Disclaimer

The study has used data from the Cancer Registry of Norway, the Norwegian Patient Registry and the Norwegian Prescribed Drug Registry. The interpretation and reporting of these data are the sole responsibility of the authors, and no endorsement by these registries is intended nor should be inferred.

## Secure data environment

De-identified individual-level data were stored, and all analyses performed on the TSD (Tjeneste for Sensitive Data) facilities, owned by the University of Oslo, operated and developed by the TSD service group at the University of Oslo, IT-Department (USIT).

## Data availability

Data extraction is available upon separate request and approval by the Norwegian Regional Committees for Medical and Health Research Ethics and the data permit authority at the Norwegian Institute of Public Health.

## 7 Author contributions

Børø, Brant, Helland, Jonasson, and Thoresen wrote the manuscript. Jonasson, Thoresen, and Brant designed the research. Brant analysed the data. All authors reviewed the manuscript.

